# Pleiotropic effects of heterozygosity for the *SERPINA1* Z allele in the UK Biobank

**DOI:** 10.1101/2020.06.04.20115923

**Authors:** Katherine A Fawcett, Kijoung Song, Guoqing Qian, Aliki-Eleni Farmaki, Richard Packer, Catherine John, Nick Shrine, Raquel Granell, Sue Ring, Nicholas J Timpson, Laura M Yerges-Armstrong, Richard Eastell, Louise V Wain, Robert A Scott, Martin D Tobin, Ian P Hall

## Abstract

Homozygosity for the *SERPINA1* Z allele causes alpha-1 antitrypsin deficiency, a rare condition that can cause lung and liver disease. However, the effects of Z allele heterozygosity on non-respiratory phenotypes, and on lung function in the general population, remain unclear. We conducted the largest population-based study to date to determine Z allele effects on >2,400 phenotypes using the UK Biobank study (N>303,353). We detected strong associations between heterozygosity and non-respiratory phenotypes including increased height, increased risk of gall bladder disease, reduced risk of heart disease and lower blood pressure, reduced risk of osteoarthritis and reduced bone mineral density, increased risk of headache and enlarged prostate, as well as with blood biomarkers of liver function. Heterozygosity was associated with higher lung function in non-smokers, but smoking appears to abolish this protective effect. Individuals heterozygous for the Z allele may therefore have altered risk of smoking-induced lung disease and other, non-respiratory conditions.

## Introduction

Homozygosity for the *SERPINA1* Z allele (rs28929474(T)) is the commonest cause of severe alpha-1 antitrypsin deficiency (AATD) and is a well-established genetic risk factor for lung diseases such as chronic obstructive pulmonary disease (COPD). However, the health consequences of heterozygosity for the Z allele are not as well-understood^1^. Given that approximately one in 30 Europeans are heterozygous for the Z allele, the phenotypic consequences of carriage of this allele could have important public health implications.

Some previous studies have sought to characterise the effect of Z allele heterozygosity and homozygosity on non-respiratory traits, particular liver diseases ^2, 3, 4, 5, 6^. However, these have often been carried out in small sample sizes and/or clinical subgroups. To date there has been no systematic approach to determine the consequences of Z allele heterozygosity and homozygosity to non-respiratory phenotypes. The recent development of phenome-wide association testing (PheWAS) and the availability of the well-phenotyped UK Biobank population-based cohort provides a platform to allow this question to be addressed.

In contrast, the effect of Z allele heterozygosity and homozygosity on lung function traits and lung disease has been the subject of many studies. Recent COPD case-control and family-based studies have shown reduced lung function and increased risk of COPD in heterozygous current and former smokers^7, 8, 9, 10^. However, a population-based study demonstrated no significant reductions in lung function in heterozygous smokers, despite having greater numbers of heterozygous smokers compared to previous studies^11^. This discrepancy may be due, in part, to the fact that study participants were identified and recruited based on their health status (as in case-control studies^7, 8^) or based on the health status of a family member^9, 10^, and this can lead to causal estimates that are subject to ascertainment bias^12^. Population-based studies^11, 13^ overcome these biases. In addition, the effects of the Z allele on lung function in relation to smoking status remain uncertain.

We therefore systematically evaluated the effects of Z allele heterozygosity in the UK biobank population, which in total includes >18,000 Z allele heterozygotes. We aimed to (i) undertake the first phenome-wide association study for Z allele heterozygosity and homozygosity to identify effects beyond the respiratory system, and (ii) to fully define the effects of Z allele heterozygosity on lung function measures (FEV_1_ and FVC) in smokers and non-smokers.

## Methods

### Cohorts

#### UK Biobank

The UK Biobank data resource is described elsewhere (see http://www.ukbiobank.ac.uk) and has ethical approval from the UK National Health Service (NHS) National Research Ethics Service (Ref 11/NW/0382). All participants provided written informed consent. Genotyping was undertaken using the Affymetrix Axiom® UK BiLEVE and UK Biobank arrays^14^. Genotypes were imputed based on the Human Reference Consortium (HRC) panel, as described elsewhere^15^. Genotyping quality control was performed as described previously^16^. UK Biobank individuals were selected for inclusion in PheWAS analyses if they met the following criteria: (i) they had no missing data for sex or age; (ii) they had genome-wide imputed genetic data for rs28929474; (iii) they were of genetically determined European ancestry and; (iv) they were not first- or second-degree relatives. For lung function analyses, individuals were additionally required to have spirometry data that passed quality control, as described previously^16^ and to have full data for height and smoking status as well as data derived from direct genotyping for rs28929474 (N = 303,353).

#### ALSPAC

ALSPAC children were genotyped using the Illumina HumanHap550 quad genome-wide SNP genotyping platform (Illumina Inc., San Diego, CA, USA) by the Wellcome Trust Sanger Institute (WTSI; Cambridge, UK) and the Laboratory Corporation of America (LCA, Burlington, NC, USA), using support from 23andMe. Ethical approval for the study was obtained from the ALSPAC Ethics and Law Committee and the Local Research Ethics Committees. Informed consent for the use of data collected via questionnaires and clinics was obtained from participants following the recommendations of the ALSPAC Ethics and Law Committee at the time. Further details of ALSPAC are available in the Supplementary materials. Participants were excluded from this study if they had incorrectly recorded sex, minimal or excessive heterozygosity, disproportionate levels of individual missingness (>3%), evidence of cryptic relatedness or non-European ancestry.

#### Phenome-wide association studies (PheWAS)

To identify whether *SERPINA1* Z allele heterozygosity or homozygosity was associated with respiratory and non-respiratory traits and diseases, two PheWAS across all available traits were performed. The first compared traits in individuals heterozygous for the Z allele vs wild-type, and the second compared individuals homozygous for the Z allele vs wild type and heterozygotes combined (recessive genetic model). Up to 379,101 individuals were available for these analyses. Traits included UK Biobank baseline measures (from both questionnaires and physical measures), selfreported medication usage, and operative procedures, as well as those captured in Office of Population Censuses and Surveys codes from the electronic health record. We also included selfreported disease variables and those from hospital episode statistics (ICD-10 codes truncated to three-character codes and combined in block and chapter groups) as well as combining both selfreport and hospital diagnosed diseases where possible to maximise power. The analysis included 2,411 traits (traits with >200 cases were included). Analyses were conducted in unrelated European-ancestry individuals (KING kinship coefficient of <0.0442), and were adjusted for age, age^2^, sex, genotyping array, and ten ancestry principal components. Logistic models were fitted for binary outcomes, and linear models were fitted for quantitative outcomes (rank transformed to normality). False discovery rates were calculated according to the number of the traits in the analysis.

#### Statistical analyses for biomarkers, lung function, COPD and height

For continuous traits, linear regression models adjusted for sex, age, age^2^, the first ten ancestry-based principal components (PCs) and genotyping array were tested in R. *SERPINA1* Z genotype (derived from direct genotyping) was coded according to the genetic model tested: either heterozygous (ie. heterozygous versus wild-type) or recessive. For lung function analyses, ever-smoking status and standing height were also included in the models unless indicated. To test for interaction of Z allele hetero-or homo-zygosity with ever-smoking status and sex, interaction terms for sex and smoking were added to the model. Association testing with moderate-severe COPD (defined as FEV_1_/FVC<0.7 and FEV_1_ < 80% predicted) was carried out using logistic regression in R adjusting for covariates sex, age, age^2^, ever-smoking status, height, the first ten ancestry-based principal components (PCs) and genotyping array. In addition to the biomarkers available from UK Biobank, we calculated estimated glomerular filtration rate (eGFR) using the following formula: If cystatin C (cys) <= 0.8 then eGFR = 133*((cys/0.8)**-0.499)*(0.996**age)*[0.932 if female] whereas if cystatin C (cys) > 0.8 then eGFR = 133*((cys/0.8)**-1.328)*(0.996**age)*[0.932 if female].

In ALSPAC, the association of Z allele heterozygosity with height and lung function z-scores were examined using linear regression models including age and gender (and including height in the lung function analyses).

#### Gene Expression

We looked for human tissue-specific expression of SERPINA1 mRNA and protein using Human Protein Atlas (https://www.proteinatlas.org/), which integrates quantitative transcriptomics and antibody-based proteomics of human tissues^17^. We interrogated potential eQTL signals in Gtex (https://gtexportal.org).

## Results

### Frequency of the *SERPINA1* Z in UK Biobank

Amongst 411,002 unrelated, European UK Biobank participants with full sex, age, height, smoking status and genotyping data, 16,199 (3.94%) were heterozygous and 129 (0.0314%) were homozygous for the Z allele. There were fewer ZZ homozygotes than expected under Hardy-Weinberg Equilibrium in all samples (P = 0.004), ever-smokers (P = 0.003), individuals over 60 years of age (P = 0.007) and in females (P = 0.007) (Supplementary Table 1), consistent with reduced survival.

### Phenome-wide association studies of the Z allele

To explore the effect of Z allele heterozygosity and homozygosity on a broad range of phenotypes we performed two PheWAS. The phenotypes most strongly associated with Z allele heterozygosity and Z allele homozygosity (FDR < 0.01) are shown in **Fig. 1A** and **1B** respectively (full results are in **Supplementary Tables 2** and **3**). Standing height, alpha 1 antitrypsin deficiency, and various respiratory phenotypes (lung function, reported emphysema, chronic bronchitis and COPD) were strongly associated with the Z allele under either genetic model. However, we also detected strong associations between Z allele heterozygosity and a number of other phenotypes, including increased risk of gall stones (OR = 1.35, P = 1.69×10^-16^), gall bladder removal (OR = 1.51, P = 1.85×10^-29^) and bile duct disease (OR = 1.25, P = 5.42×10^-8^); cardiovascular phenotypes such as lower risk of heart disease (OR = 0.84, P = 6.66×10^-6^) and family history of heart disease, and lower blood pressure (P = 1.02×10^-10^); musculoskeletal phenotypes such as a higher grip strength, lower risk of osteoarthritis, particularly of knee or hip (OR = 0.85, P = 1.18×10^-6^), lower bone mineral density (P = 1.40×10^-5^) and higher risk of osteoporosis (OR = 1.17, P = 0.001); and also higher risk of headache (OR = 1.12, P = 5.66×10^-9^) and enlarged prostate (OR = 1.26, P = 1.95×10^-7^). The pheWAS also included blood counts for a variety of different cell types, showing reduced reticulocytes in individuals heterozygous for the Z allele. Z allele homozygosity was associated with increased haemoglobin (P = 7.17×10^-10^), haematocrit (P = 6.17×10^-8^) and red blood cells (P = 5.91×10^-5^), as well as optic neuritis (OR = 26.9, P = 4.32×10^-6^) and pancreatitis (OR = 6.07, P = 7.76×10^-5^).

**Fig. 1.**
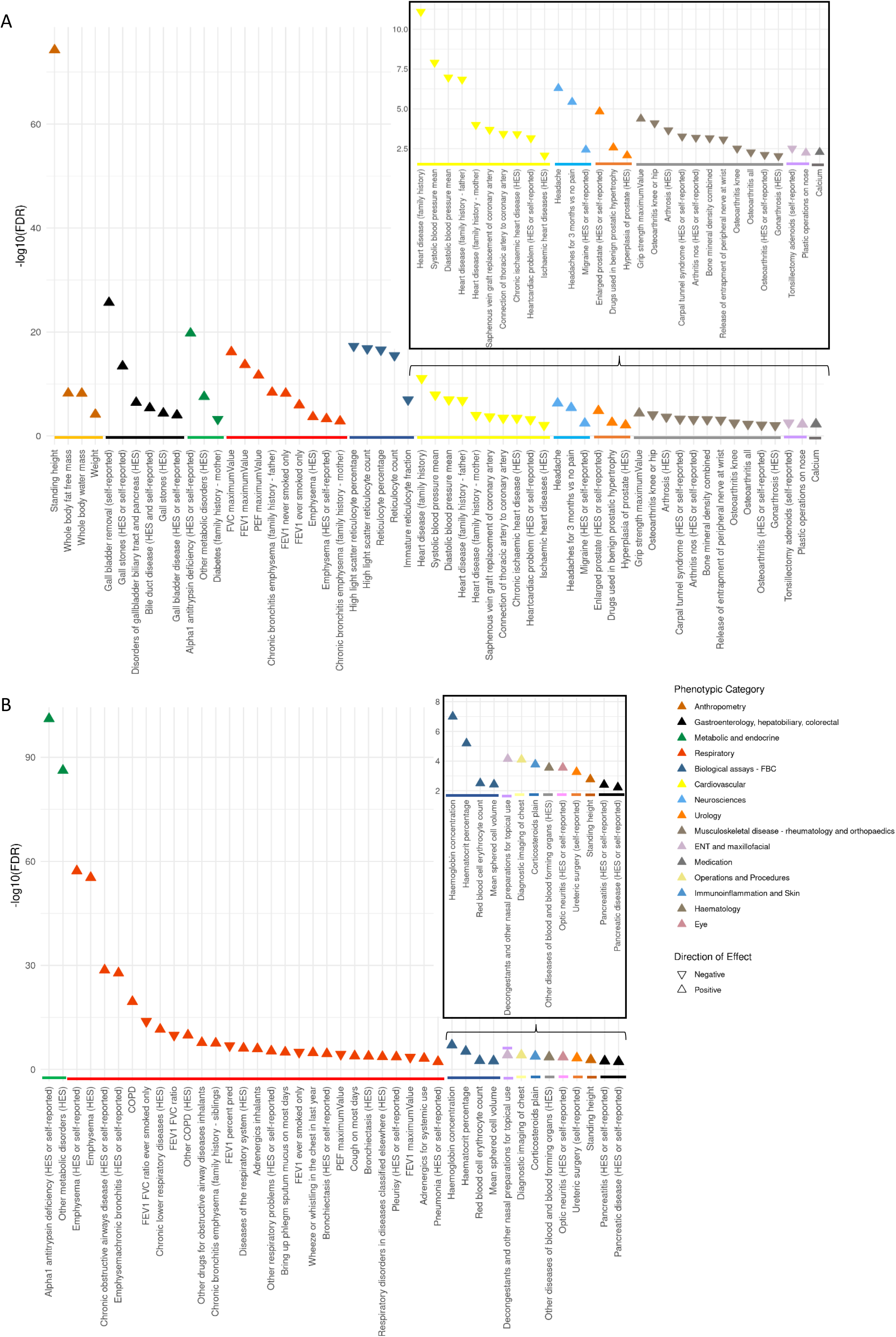
Traits associated with the *SERPINA1* Z allele in UK Biobank. Traits displayed are those with a false discovery rate<0.01 from phenome-wide association study of 2411 phenotypes across 379101 UK Biobank participants using the heterozygous model for the Z allele (wild-type vs heterozygous individuals) (A) and the recessive model for the Z allele (wild-type and heterozygous individuals vs individuals homozygous for the Z allele) (B).

To extend these analyses to traits not previously available for the PheWAS, we investigated the effect of Z allele heterozygosity and homozygosity on thirty key blood biochemistry markers in UK Biobank data (**Table 1**). Heterozygosity for the Z allele was associated with a variety of liver function markers including higher alanine aminotransferase (β = 1.16 U/l, P = 2.95×10^-25^), higher albumin (P = 0.71 g/l, P = 3.11×10^-229^), higher alkaline phosphatase (β = 2.97 U/l, P = 5.81×10^-44^), higher aspartate aminotransferase (0.71 U/l, P = 2.27×10^-16^) and higher direct bilirubin (β = 0.041 umol/l, P = 1.64×10^-8^). There were also strong associations with calcium (β = 0.012 mmol/l, P = 4.01×10^-50^), cystatin C (0.0090 mg/l, P = 8.91×10^-12^), C-reactive protein (β = −0.17 mg/l, P = 1.11×10^-6^) and IGF-1 (β = −0.21 nmol/l, P = 2.35×10^-6^). Heterozygotes exhibited higher sex hormone binding globulin (SHBG) (β = 4.53 nmol/l, P = 1.06×10^-95^) and testosterone (β = 0.37 nmol/l, P = 1.84×10^-57^). The association with higher testosterone was driven by heterozygosity in males (0.76 nmol/l [SE 0.045], P = 3.38×10^-64^), but not females (−0.030 nmol/l [SE 0.0079], P = 0.00015).

**Table 1.** Association between the *SERPINA1* Z allele and blood biomarkers in unrelated, European UK Biobank participants

| Biomarker | Heterozygous genetic model |  | Recessive genetic model |  |
| --- | --- | --- | --- | --- |
|  | Beta (se) | p | Beta (se) | p |
| Alanine aminotransferase (U/l) | 1.163 (0.112) | **2.95x10<sup>-25</sup>** | 3.037 (1.246) | 0.015 |
| Albumin (g/l) | 0.712 (0.022) | **3.11x10<sup>-229</sup>** | 0.306 (0.243) | 0.208 |
| Alkaline phosphatase (U/l) | 2.968 (0.213) | **5.81x10<sup>-44</sup>** | 2.379 (2.373) | 0.316 |
| Apolipoprotein B (g/l) | -0.0037 (0.0021) | 0.082 | 0.042 (0.024) | 0.086 |
| Apolipoprotein A (g/l) | -0.0045 (0.0019) | 0.020 | 0.019 (0.022) | 0.375 |
| Aspartate aminotransferase (U/l) | 0.707 (0.086) | **2.27x10<sup>-16</sup>** | 6.353 (0.956) | **3.04x10<sup>-11</sup>** |
| C-reactive protein (mg/l) | -0.174 (0.036) | **1.11x10<sup>-6</sup>** | 0.192 (0.396) | 0.628 |
| Calcium (mmol/l) | 0.012 (0.00080) | **4.01x10<sup>-50</sup>** | 0.01 (0.009) | 0.265 |
| Cholesterol (mmol/l) | 0.0051 (0.0092) | 0.576 | 0.19 (0.102) | 0.063 |
| Creatinine (umol/l) | -0.067 (0.129) | 0.601 | -0.94 (1.435) | 0.512 |
| Cystatin C (mg/l) | 0.0090 (0.0013) | **8.91x10<sup>-12</sup>** | 0.019 (0.015) | 0.204 |
| Direct bilirubin (umol/l) | 0.041 (0.0073) | **1.64x10<sup>-8</sup>** | 0.158 (0.079) | 0.046 |
| eGFR* | -0.241 (0.157) | 0.125 | -1.810 (1.732) | 0.296 |
| Gamma glutamyltransferase (U/l) | 0.253 (0.342) | 0.460 | 8.045 (3.818) | 0.035 |
| Glucose (mmol/l) | -0.0032 (0.010) | 0.758 | -0.133 (0.114) | 0.244 |
| Glycated haemoglobin (HbA1c) (mmol/mol) | -0.187 (0.052) | **3.40x10<sup>-4</sup>** | -1.781 (0.588) | 2.46x10<sup>-3</sup> |
| HDL cholesterol (mmol/l) | 0.0015 (0.0030) | 0.605 | 0.142 (0.033) | **2.01x10<sup>-5</sup>** |
| IGF-1 (nmol/l) | -0.212 (0.045) | **2.35x10<sup>-6</sup>** | -2.245 (0.5) | **7.19x10<sup>-6</sup>** |
| LDL direct (mmol/l) | 0.011 (0.007) | 0.113 | 0.115 (0.079) | 0.145 |
| Lipoprotein A (nmol/l) | -0.324 (0.452) | 0.474 | -0.151 (5.115) | 0.976 |
| Oestradiol (pmol/l) | 9.577 (8.441) | 0.257 | 88.051 (96.144) | 0.360 |
| Phosphate (mmol/l) | 0.0027 (0.0013) | 0.042 | -0.014 (0.015) | 0.352 |
| Rheumatoid factor (IU/ml) | 0.113 (0.529) | 0.832 | -3.27 (5.7) | 0.566 |
| SHBG (nmol/l) | 4.531 (0.218) | **1.06x10<sup>-95</sup>** | 40.573 (2.402) | **5.33x10<sup>-64</sup>** |
| Testosterone (nmol/l) | 0.37 (0.023) | **1.84x10<sup>-57</sup>** | 2.986 (0.258) | **6.96x10<sup>-31</sup>** |
| Total bilirubin (umol/l) | 0.146 (0.035) | **3.54x10<sup>-5</sup>** | 0.52 (0.391) | 0.184 |
| Total protein (g/l) | 0.578 (0.035) | **1.06x10<sup>-62</sup>** | 0.459 (0.382) | 0.229 |
| Triglycerides (mmol/l) | -0.023 (0.0082) | 4.69x10<sup>-3</sup> | -0.324 (0.092) | **4.21x10<sup>-4</sup>** |
| Urate (umol/l) | 1.096 (0.56) | 0.050 | -1.325 (6.252) | 0.832 |
| Urea (mmol/l) | 0.051 (0.011) | **2.69x10<sup>-6</sup>** | 0.06 (0.122) | 0.620 |
| Vitamin D (nmol/l) | -0.041 (0.175) | 0.816 | -2.547 (1.955) | 0.193 |

Results are based on linear regression adjusting for age, age<sup>2</sup>, ancestry-based principal components,
and genotyping array. The additive or recessive genetic model was also included in the regression as
shown. Highlighted in bold are P values significant at the Bonferroni-corrected level

### Z allele heterozygosity is associated with increased lung function in non-smokers

We tested association of heterozygosity for the *SERPINA1* Z allele with lung function traits in unrelated, European UK Biobank participants with full sex, age, height, smoking status, lung function and genotyping data (**Table 2**). Individuals heterozygous for the Z allele exhibited higher FEV_1_ (β = 9.26 ml, P = 0.041) but no association with FEV_1_/FVC or FVC compared to wild-type (**Table 2**). However, in stratified analyses of UK Biobank never- and ever-smokers, we found that heterozygosity for the Z allele was associated with a large increase in FEV_1_ (β = 19.36 ml, P = 9.21×10^-4^) and increased FEV_1_/FVC (β = 0.0031, P = 1.22×10^-5^) in never-smokers, but not in ever-smokers (Table 2). Statistical tests of Z allele*ever-smoking interactions showed interactions for FEV_1_ (P = 0.022) and FEV_1_/FVC (P = 1.06×10^-4^) (**Supplementary Table 4**). Furthermore, heterozygous ever-smokers exhibited a small increased risk of COPD (OR = 1.16 [95% CI 1.05 – 1.29], P = 0.005), whereas heterozygous never-smokers did not (OR = 0.87 [95% CI 0.75 – 1.01], P = 0.070) **(Table 2**).

**Table 2.** Association between the *SERPINA1* Z allele and height, lung function traits, and COPD in unrelated, European UK Biobank participants

| Genetic model | Smoking status | N (test vs comparison) | Height (cm) |  | FEV<sub>1</sub> (ml) |  | FEV<sub>1</sub>/FVC |  | FVC (ml) |  | COPD* |  |
| --- | --- | --- | --- | --- | --- | --- | --- | --- | --- | --- | --- | --- |
|  |  |  | Beta (se) | p | Beta (se) | p | Beta (se) | p | Beta (se) | p | OR (95% CI) | P |
| Het | All | 11877 vs 291383 | 1.023<br>(0.059) | **3.91x10<sup>-68</sup>** | 9.26<br>(4.53) | **0.041** | 0.00091<br>(0.00057) | 0.108 | 9.38<br>(5.43) | 0.084 | 1.05 (0.96-1.14) | 0.268 |
|  | Ever-smokers | 5357 vs 133779 | 1.027<br>(0.087) | **8.53x10<sup>-32</sup>** | -2.63<br>(7.05) | 0.709 | -0.0017<br>(0.00091) | 0.062 | 5.47<br>(8.28) | 0.509 | 1.16 (1.05-1.29) | **0.005** |
|  | Never-smokers | 6520 vs 157604 | 1.022<br>(0.079) | **3.03x10<sup>-38</sup>** | 19.36<br>(5.84) | **9.21x10<sup>-4</sup>** | 0.0031<br>(0.00071) | **1.22x10<sup>-5</sup>** | 12.77<br>(7.15) | 0.074 | 0.87 (0.75-1.01) | 0.070 |
| Rec | All | 93 vs 303260 | 1.644<br>(0.650) | **0.011** | -249.21<br>(50.22) | **6.96x10<sup>-7</sup>** | -0.051<br>(0.0063) | **3.54x10<sup>-16</sup>** | -132.16<br>(60.09) | **0.028** | 7.42 (4.29 - 12.27) | **5.67x10<sup>-14</sup>** |
|  | Ever-smokers | 35 vs 139136 | 1.242<br>(1.062) | **0.242** | -379.21<br>(85.54) | **9.30x10<sup>-6</sup>** | -0.081<br>(0.011) | **2.00x10<sup>-13</sup>** | -177.95<br>(100.35) | **0.076** | 9.19 (4.19-19.00) | **6.27x10<sup>-9</sup>** |
|  | Never-smokers | 58 vs 164124 | 1.873<br>(0.821) | **0.023** | -174.68<br>(60.68) | **0.004** | -0.034<br>(0.0074) | **4.95x10<sup>-6</sup>** | -107.69<br>(74.23) | 0.147 | 5.98 (2.66-12.03) | **2.69x10<sup>-6</sup>** |

Results are based on linear (or logistic\*) regression adjusting for age, age<sup>2</sup>, ancestry-based principal components, genotyping array and, in the case of lung function and COPD, standing height. The heterozygote or recessive genetic model was also included in the regression as shown. COPD was spirometrically defined as FEV<sub>1</sub>/FVC<0.7 and FEV<sub>1</sub> < 80% predicted (consistent with spirometric definition of GOLD Stage 2-4)

### The Z allele is strongly associated with height

Heterozygosity for the Z allele was strongly associated with height (β = 1.02 cm, P = 3.91×10^-68^ – Table 2) and, when lung function measures were not adjusted for height, the Z allele heterozygosity had much larger effect estimates for FEV_1_ (β = 44.72 ml [SE 4.97], P = 2.20×10^-19^) and FVC (β = 61.06 ml [SE 6.18], P = 5.18×10^-23^), but not FEV_1_/FVC. These results suggest that carrying one copy of the Z allele confers an advantage primarily (but not entirely) driven by increased height. To investigate the developmental stage at which Z allele heterozygosity influences height, we tested association with height in the ALPSAC cohort at ages 8, 15 and 24 years. The findings were consistent with Z allele heterozygosity influencing height from adolescence and, at all the ages, directions of effect on height and lung function were consistent with those in UK Biobank (**Supplementary Table 5**).

### Sex modifies the effect of Z allele heterozygosity on lung function

We also assessed whether sex can modify the effect of the Z allele on lung function or height and found that heterozygosity for the Z allele was associated with higher height-adjusted FEV_1_/FVC in females (β = 0.0028 [SE 0.00071], P = 5.89×10^-5^) but not in males (β = −0.0015 [SE 0.0009], P = 0.093) (Z allele*sex interaction: P = 0.00021 for FEV_1_/FVC, **Supplementary Table 4** and **Supplementary Table 6**).

### Association between Z allele homozygosity and lung function

In contrast to the lung function raising effects of carrying one copy of the Z allele, homozygosity for the Z allele has been widely reported to reduce lung function and increase risk of COPD. Similarly, UK Biobank individuals homozygous for the Z allele had significantly lower FEV_1_ (β = −249.21 ml, P = 6.96×10^-7^), FEV_1_/FVC (β = −0.051, P = 3.54×10^-16^) and FVC (β = −132.16, P = 0.028) compared to wild-type and heterozygous individuals and greater risk of COPD (OR = 7.42, P = 5.67×10^-14^) (**Table 2**). The associations between homozygosity for the Z allele and reduced lung function and increased risk of COPD were stronger in smokers compared to non-smokers (**Table 2**). Given the strength of these associations, it is interesting to note that 67 of 93 individuals homozygous for the Z allele do not have spirometrically-defined COPD and eight out of 17 homozygous ever-smokers aged over 60 do not have spirometrically-defined COPD. We generated polygenic risk scores for lower FEV_1_/FVC by weighting variants according to beta-coefficients in the SpiroMeta consortium cohorts^16^. The 67 individuals homozygous for the Z allele but without COPD have an average score percentile of 48.4 compared to 58.2 in those with COPD. This suggests that individuals homozygous for the Z allele but without COPD may have a more protective genetic profile across other genomic loci.

### Diagnosis of AATD in individuals homozygous for the Z allele

We note that only 20 of 141 individuals who were genotyped ZZ homozygotes in the full UK Biobank cohort had a recorded ICD-10 coding for AATD (E880) in Hospital Episode Statistics. As this could indicate miscoding rather than misdiagnosis, we investigated further in subset of 65 homozygotes with linked primary care data. Of these 65 ZZ homozygotes, 15 had primary care (Read) codes or ICD-10 codes for AATD and 50 did not. Of the group without AATD codes, four had primary care (Read) codes or ICD-10 codes for bronchiectasis and an additional 6 had primary care (Read) codes or ICD-10 codes for COPD indicating potential misdiagnosis of AATD.

### *SERPINA1* expression profiles

Interrogation of data on *SERPINA1* expression showed strongest expression both at the mRNA and protein level in liver. Expression was also seen in lung, stomach, smooth muscle, small bowel, testis and kidney, although expression levels were generally low and somewhat variable between different contributing datasets. SNP rs28929474 is an eQTL signal for the IFI27L2 (interferon alpha inducible protein 27 like 2), in left ventricle and artery, with Z allele heterozygotes showing reduced expression compared with wild type.

## Discussion

In the largest population-based study to date we describe novel phenotypic associations of heterozygosity for the Z allele of SERPINA1 and also present definitive evidence that heterozygosity for the Z allele is associated with greater lung function only in non-smokers. Our findings have implications for individuals heterozygous for the *SERPINA1* Z allele, and more generally for the study of rare variants and gene-environment interactions.

Our PheWAS examining the consequences of carrying one copy of the SERPINA1 Z allele revealed potentially important associations with non-respiratory traits and diseases. Consistent with previous reports^11^, we detected a very strong association between Z allele heterozygosity and height (with each additional Z allele adding approximately 1cm to height). The mechanism and timing of the effect of the Z allele on height is not understood. Our findings in the ALSPAC study suggest a possible influence of Z allele heterozygosity on height which is manifest from puberty, although results should be interpreted with caution given the limitations in power in ALSPAC alone. This suggests that sex hormones are a possible mediator of the height-raising effects of the Z allele and, indeed, we detected higher levels of testosterone in individuals heterozygous for the Z allele. However, heterozygosity for the Z allele increases height in males and females, whereas increased testosterone is only seen in males. Furthermore, Z allele heterozygotes also exhibit higher SHBG suggesting that the amount of free testosterone will be unchanged. IGF-I levels relate to height, but in our study the magnitude of the association was very small (less than 1% average levels) and it was a negative association. It is therefore still not clear why heterozygosity and homozygosity for the Z allele increases height.

We detected strong associations between heterozygosity for the Z allele and increased risk of gall stones and gall bladder removal. This is consistent with recent reports that the Z allele may be a risk factor for developing gallstone disease^18, 19^. The mechanism underlying this association is unclear but previous reports have suggested it may be related to liver dysfunction in AATD and/or the composition of bile^18, 19^. We found that individuals heterozygous for the Z allele also had increased levels of biomarkers such as alanine aminotransferase, alkaline phosphatase, aspartate aminotransferase and bilirubin, consistent with an impact of heterozygosity on liver function.

Heterozygosity for the Z allele was also associated with musculoskeletal phenotypes such as reduced risk of osteoarthritis, increased risk of osteoporosis and lower bone mineral density. Calcium was increased in heterozygotes but as albumin was also increased free calcium may not be different in this group. A recent study showed that AAT can inhibit receptor activator of nuclear factor κ-B ligand (RANKL)-induced osteoclast formation and bone resorption^20^. As heterozygosity for the Z allele causes reduction in AAT, individuals heterozygous for the Z allele may have increased risk of osteoporosis and lower bone mineral density due to increased RANKL-induced bone resorption. Lower bone mineral density has been associated with a lower risk of osteoarthritis^21, 22^.

Heterozygotes for the Z allele also had reduced risk of heart disease (even when adjusted for height) and lower blood pressure, as well as increased risk of headache consistent with one previous report suggesting cluster headaches may be more frequent in heterozygotes for the Z allele^6^. There was also a marked association between heterozygosity and reduced reticulocyte count and reticulocyte percentage which, as far as we are aware, has not been reported previously.

Homozygosity for the Z allele was strongly associated with alpha 1 antitrypsin deficiency and respiratory traits, but also haemoglobin concentration and haematocrit percentage, which may be driven by the associations with respiratory disease. Homozygosity was also associated with markedly higher risk of optic neuritis and pancreatitis. Whilst the association with pancreatitis could be driven by the increased rick of gall stones, the mechanism underlying the increased risk of optic neuritis is unclear.

Whilst homozygosity for the Z allele is an established cause of respiratory disease, the effect of heterozygosity on respiratory health of general populations has been less clear. Our study provides definitive data on the effects of heterozygosity for the Z allele on respiratory traits, showing that heterozygosity is associated with greater lung function in never-smokers (even after height adjustment), but not in smokers. Our results contrast with previous studies^7, 8, 9, 10^ that did not detect higher lung function in participants heterozygous for the Z allele compared to wild-type, and with studies that did not detect differential effects by smoking status^11^. There are several possible reasons for these discrepancies. First, the relatively low frequency of the Z allele meant previous studies were underpowered. Our study contained 18 times more heterozygous participants than the previous largest population-based study and 46 times more than the largest case-control study. Whilst one previous study has suggested a beneficial effect of Z allele heterozygosity on lung function^11^ it nevertheless did not detect differential effects by smoking status. Again, this is probably due to power. Second, most of the previous studies were conducted in current and former smokers only^7, 8, 10^, and would therefore have missed the effect of Z allele heterozygosity in nonsmokers. Third, most previous studies recruited participants on the basis of their health status (as in case-control studies^7, 8^) or the health status of a family member^9, 10, 23^. These approaches can lead to ascertainment biases that might distort causal estimates. A major strength of using the UK Biobank for our study was that participants were not ascertained on the basis of having lung disease, enabling a relatively unbiased assessment of the role of the heterozygosity for the Z allele in lung function. Fourth, the Z allele is not present on most standard genotyping arrays, nor are there any adequate proxies^11^ - issues that disproportionately affect low frequency and rare variants. Furthermore, genetic studies tend to focus on the additive genetic model rather than heterozygous or recessive models. Because we designed the Z allele into the UK Biobank array^14^ and compared different genetic models, we were able to robustly examine this association in UK Biobank participants.

The biological mechanism underlying the association between heterozygosity for the Z allele and increased lung function in non-smokers is not clear. The *SERPINA1* gene encodes alpha 1 antitrypsin (AAT), an enzyme with an important role in inhibiting proteases, such as neutrophil elastase, which are secreted during inflammation and cause collateral tissue damage. The Z allele produces AAT with reduced anti-protease activity and a propensity to form polymers, which accumulate in liver thereby reducing the circulating levels of AAT. The polymers themselves have also been shown to be a pro-inflammatory stimulus in lung tissue^24^ which further perpetuates lung inflammation and damage. Homozygosity for the Z allele is the most common cause of severe AAT deficiency whereas heterozygosity for the Z allele has been shown to result in intermediate levels of AAT (~60% of normal)^25^. One might therefore expect heterozygosity to have, if anything, a detrimental effect on lung function. There is evidence that positive selection has acted at the Z allele locus^11, 26^ and mechanisms of heterozygous advantage (through increased height^11^ and protection from infectious respiratory diseases by promotion of inflammatory responses^26^) have been proposed. The amplification of inflammatory response as a result of cigarette smoke may therefore provide a mechanism for eliminating this advantage in smokers. It has also been shown that smoking-induced oxidative modifications to AAT reduce its ability to inhibit neutrophil elastase and turn it into a pro-inflammatory mediator^27^. Cigarette smoke has been found to accelerate polymerisation of Z-type AAT^28^, suggesting a possible mechanism whereby smoking may modify the effect of the Z allele on lung function.

We also report a novel interaction between sex and the *SERPINA1* Z allele heterozygosity on FEV_1_/FVC. Women heterozygous for the Z allele exhibited greater FEV_1_/FVC compared to wild-type, whereas men heterozygous for the Z allele showed no significant difference to wild-type. Previous studies have shown that individuals with the fastest decline in lung function amongst AATD patients are more likely to be male^29^. However, the reasons why sex would appear to modify the effect of Z allele heterozygosity on lung function are not clear.

In conclusion, we have used by far the largest and best-powered study to date to demonstrate that heterozygosity for the Z allele is associated with previously unrecognised non-respiratory phenotypes, and also with greater lung function in non-smokers. Our results demonstrate that large sample sizes are required to study the effects of rare variants, particularly when these effects depend on an environmental stimulus. The opposing directions of effect of Z allele heterozygosity and homozygosity demonstrate that the additive genetic model is not always appropriate for genetic studies of the rarer variants such as the *SERPINA1* Z allele. Our findings suggest that while individuals who are heterozygous for the Z allele may not exhibit symptoms of AAT deficiency, they may be more susceptible to smoking-induced lung disease and have altered risk of other non-respiratory conditions.

## Data Availability

All the results of this study are available in the main manuscript and supplementary material.

## Acknowledgements

The UK Biobank analysis was conducted under approved UK Biobank data application number 648. We thank UK Biobank and all the participants for generating this important health research resource. This study used the ALICE and SPECTRE High Performance Computing Facilities at the University of Leicester. We are extremely grateful to all the families who took part in the ALSPAC study, the midwives for their help in recruiting them, and the whole ALSPAC team, which includes interviewers, computer and laboratory technicians, clerical workers, research scientists, volunteers, managers, receptionists and nurses.

## Author contributions

K.A.F., M.D.T., and I.P.H. designed the study, interpreted the results and wrote the manuscript. K.S., L.M.Y-A., and R.A.S. designed and conducted the PheWAS. G.Q. carried out expression analyses. K.A.F. and A-E.F. carried out the lung function analyses in UK Biobank. R.P. and C.J. contributed to the analysis of primary care data and interpretation of results. N.S. generated polygenic risk scores and contributed to study design and interpretation of results. L.V.W. contributed to study design and interpretation of results. R.G., S.R., and N.J.T. contributed the analyses in ALSPAC, and R.E. contributed to interpretation of the results. All authors reviewed the manuscript prior to submission.

